# The State of Sleep: Understanding Insomnia Symptoms and Treatments in US Adults

**DOI:** 10.1101/2025.08.21.25333428

**Authors:** Matthew Fisher, Michael L. Perlis, Andrew D. Krystal, Matthew J. Heffler

## Abstract

**Background:** Insomnia symptoms are common among US adults and there are many options for self-treatment. A wide range of prescription medicines, over the counter (OTC) options, and recreational substances are available that may be labeled for or repurposed to be used to treat insomnia symptoms.

**Methods:** A cross-sectional survey was developed and deployed via the Ipsos KnowledgePanel^®^ platform. Qualified subjects were those that reported at least 2 nights of insomnia symptoms per month or less than 2 nights of difficulty due to the use of an active treatment.

**Results:** A total of 2,223 individuals entered the survey, with 1,299 qualifying for participation as per the qualification criteria (58% prevalence rate). 1,244 participants completed the entire survey. Among those reporting recent sleep treatment use, the most used treatments were vitamin/supplement-based products, OTC allergy/cold medicines, OTC pain relief/sleep combinations, cannabis products, off-label prescription medicines, and alcohol. 74% of respondents indicated that they are not currently seeing an HCP to address their insomnia symptoms.

**Conclusion:** More than half of American adults reported suffering two or more nights per month of impaired sleep or are actively treating insomnia symptoms. A substantial number were repurposing medications or turning to marijuana/THC and alcohol in an attempt to treat these symptoms. Despite the prevalence of insomnia symptoms, few reported seeking input from an HCP. These results emphasize the widespread nature of sleep difficulties and the common nature of self-treatment. A safe and effective OTC sleep aid could potentially play an important public health role given these circumstances.

## Background

Insomnia symptoms exert a significant burden both globally and in the US. Evidence suggests that 50-70 million US adults have some kind of sleep disorder and nearly one-third of adults report some level of difficulty sleeping [1–3]. Untreated sleep disturbance is associated with increased risk for impairment across several domains, including diminished quality of life, worsening mental and physical health, and increased use of illicit substances [4, 5]. In contrast, improved sleep is associated with improvements in quality of life, performance, and both overall mental and physical health [6, 7].

The economic burden associated with untreated insomnia symptoms is also notable, with estimates suggesting that annual healthcare costs may be more than $94.9 billion [8]. Taken together, the risks and costs of sleep disturbance underscore the need for a better understanding of patient preferences and behaviors with respect to treatment. Further, whereas significant evidence supports the use of Cognitive Behavior Therapy for insomnia (CBT-I) [9–11], and it remains the first line treatment for insomnia disorder, not all patients are aware of, or elect to engage in this form of treatment [7, 12–16]. Those that are aware often do not receive CBT-I owing to a preference for medications, time concerns, financial limitations, or lack of access [17, 18]. Research also shows that the majority of patients with insomnia do not seek professional treatment through an HCP for months-to-years [19, 20], which limits patient ability to access prescription medications, leaving them more likely to opt for sleep hygeine techniques, over the counter (OTC) options, and recreational substance use.

This study sought to quantify the prevalence and impact of insomnia symptoms among the adult population in the United States by utilizing a nationally representative cross-sectional survey design. Furthermore, treatment patterns, preferences, and frequency of doctor-patient interaction regarding sleep were investigated.

## Methods

### Study Overview

To meet the study objectives, a 25-minute cross-sectional quantitative survey was designed. Respondents who met the study inclusion criteria were recruited through the Ipsos KnowledgePanel^®^ to participate in research. The subject panel is sourced from address-based sampling and rigorously maintained to provide a statistically valid representation of the United States population, making findings highly generalizable to the broader population of the country. Data was collected on topics related to sleep quality and patterns, impact of insomnia symptoms and actions taken, sleep treatment history and preferences, and healthcare provider interactions and recommendations. These data were collected via utilization of validated questionnaires and custom survey questions. Data were collected in a self-report fashion, utilizing an online survey data collection platform for data capture and storage.

The survey request was sent via email to members of the panel who have expressed a willingness to participate in research studies. The interview request included a statement of informed consent and the set of screening questions (see Inclusion Criteria and Exclusion criteria) to determine eligibility. Those who expressed interest and were eligible to participate were directed to complete a self-administered online survey. The questionnaire was completed independently on the respondent’s chosen device. The online survey could be completed at a time and location of the respondents choosing. The respondent could choose to quit the survey at any time without penalty. Respondents remained blinded to the study sponsor until all questions had been answered, at which time participants were able to request details on the study sponsor in line with local regulations. All fully completed surveys were utilized for analysis.

### Study Population and Inclusion and Exclusion Criteria

The target population for the present study were individuals that would have a propensity for treating their insomnia symptoms and for potentially seeking HCP consultation or actively engaging in treatment, defined as having difficulty falling or staying asleep 2 or more nights in the past month without medication or other substances to aid in sleep, or utilizing medication/other substances to avoid insomnia symptoms. All participants were adults currently residing in the United States.

The following inclusion criteria were used: (1) aged 18+, (2) primary or shared decision maker for own healthcare decisions, and (3) has recurrent (more than once per month) insomnia symptoms OR does not experience insomnia symptoms due to active treatment (taking a medication, supplement, or other substance) for sleep. The exclusion criteria included being unwilling to provide informed consent.

### Study Measures

The survey instrument was developed by a research team with expertise in sleep medicine as well as cross-sectional research methodologies. The survey was reviewed with a health literacy lens by researchers specializing in patient research and trained to maximize patient survey comprehension. Additionally, feedback was sought from a panel of experts in sleep medicine to further refine the survey instrument. This group included one board certified psychiatrist with specialized training in sleep medicine and extensive clinical research expertise as well as two PhD level practitioners with behavioral sleep medicine training and broad clinical and research experience. Validated measures were also leveraged along with custom survey questions.

#### Custom Survey Items

Demographic information (age, race/ethnicity, gender, marital status, education, income, work status, geographic location, comorbidities, home ownership status, household size/composition).

Sleep difficulty frequency, type of sleep difficulty found to be most bothersome, perceived importance of sleep, self-rated quality of sleep, hours of sleep per night needed to feel rested and frequency of achieving this goal, and evolution of insomnia symptoms over time. Sleep treatment awareness and experience using / trigger(s) for treatment. The survey also included an attitudinal battery relating to the impacts of insomnia symptoms.

#### Validated Measure

The Insomnia Severity Index (ISI) [21] is a 7-item instrument with item scores of 0-4 [none to severe]. The first three items pertain to the type of insomnia (initial, middle, and/or late insomnia). The remaining 4 items pertain to the daytime consequences of insomnia (e.g., <u>satisfaction</u> with current sleep ability; <u>noticeability</u> [how evident it is to others that one has a sleep problem]; <u>worry</u> [concerns about sleeplessness]; and <u>interference</u> [the extent to which daytime function is impaired owing to sleep disturbance]). Total scores range from 0-28 (with higher scores suggesting more severe insomnia). The scale score for “subthreshold” [mild] insomnia is 8-14 and clinically significant insomnia is ≥15.

#### Assessment of Insomnia Symptom Treatments

For those attempting to treat their insomnia symptoms, details about medication or substance use were obtained including obtaining prescription (if a prescription treatment was used), frequency of treatment use, self-rated improvement in sleep when utilizing treatment, actions taken the day after poor sleep, and openness to utilizing sleep treatments not currently being utilized. History of discussing insomnia symptoms with a healthcare provider (HCP) including type of HCP seen and frequency of discussing sleep with this HCP, level of satisfaction with HCP interaction and why, who initiated sleep discussions (HCP or patient), history of HCP treatment recommendation, and reasons for lack of HCP interaction for those who have not discussed insomnia symptoms with an HCP.

### Statistical Analysis

All eligible participants meeting the study inclusion criteria were included in research and analysis. While the data source for this research is representative of the American general population, the actual sample achieved for the survey deviates slightly from the overall panel composition as not every respondent on a research panel responds to all possible surveys offered by the panel. Once fieldwork was completed, data was weighted accordingly in order to achieve a statistically accurate representation of the American general population based on the weighting benchmark distributions shown in Supplementary Table 1. All analyses outlined in this paper were performed on this weighted dataset. Descriptive univariate statistics are presented for study respondents. Variables presented include the individual totals along with the respective percentages out of the total sample size for each applicable question. Calculations were performed in Microsoft Excel and SAS.

The study was conducted in accordance with legal and regulatory requirements, as well as with scientific purpose, value and rigor and follow generally accepted research practices described in Guidelines for Good Pharmacoepidemiology Practices (GPP) issued by the International Society for Pharmacoepidemiology (ISPE) and Good Practices for Outcomes Research issued by the International Society for Pharmacoeconomics and Outcomes Research (ISPOR). The study was deemed exempt according to 45 CFR 46.104(d)(2) Tests, Surveys, Interviews 45 CFR 46.104(d)(3)(i)(A) by Pearl Institutional Review Board (IRB).

## Results

In the current study, 4,319 panel members received an invitation to participate in the survey, and 2,223 potential participants entered the survey. 1,299 participants qualified for the survey based on sleep difficulty or active use of a sleep treatment, suggesting a 58% prevalence rate of difficulty with sleep in the prior month as per the study definition, as shown in Figure 1. 1,244 participants completed the survey in its entirety. Among the 1,244 total respondents, mean age was 52.5 years, and 53% were female. The majority were married (55%) and lived with a spouse or partner (65%). Please refer to Table 1. for full study demographics.

**Figure 1.**
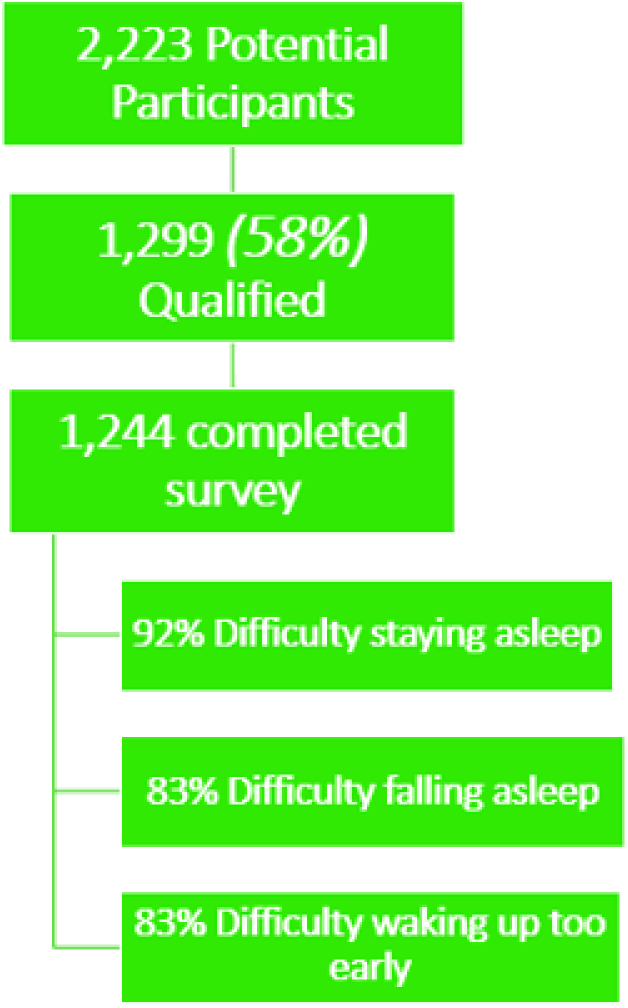
Recruitment Funnel

**Table 1.**
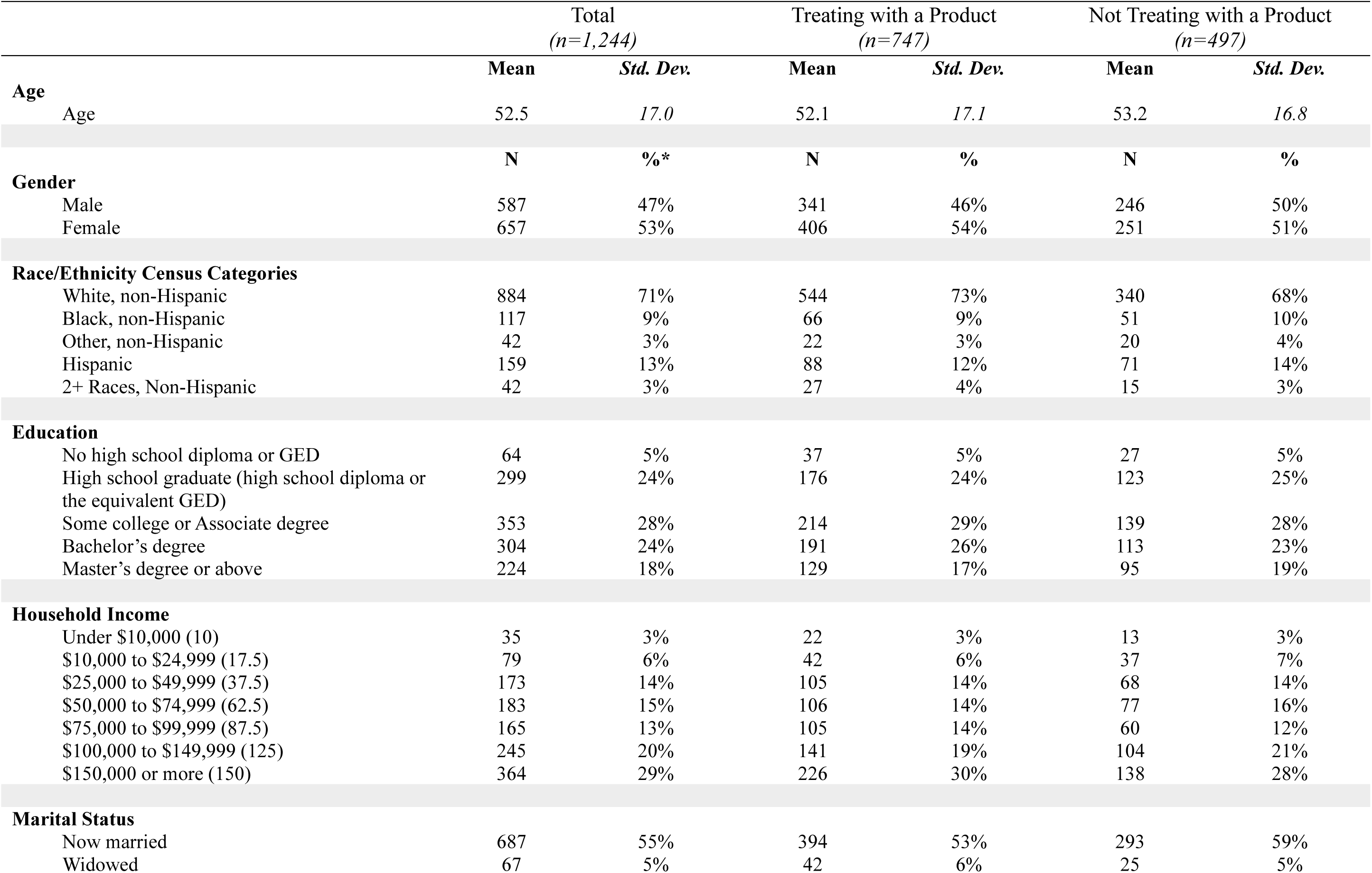
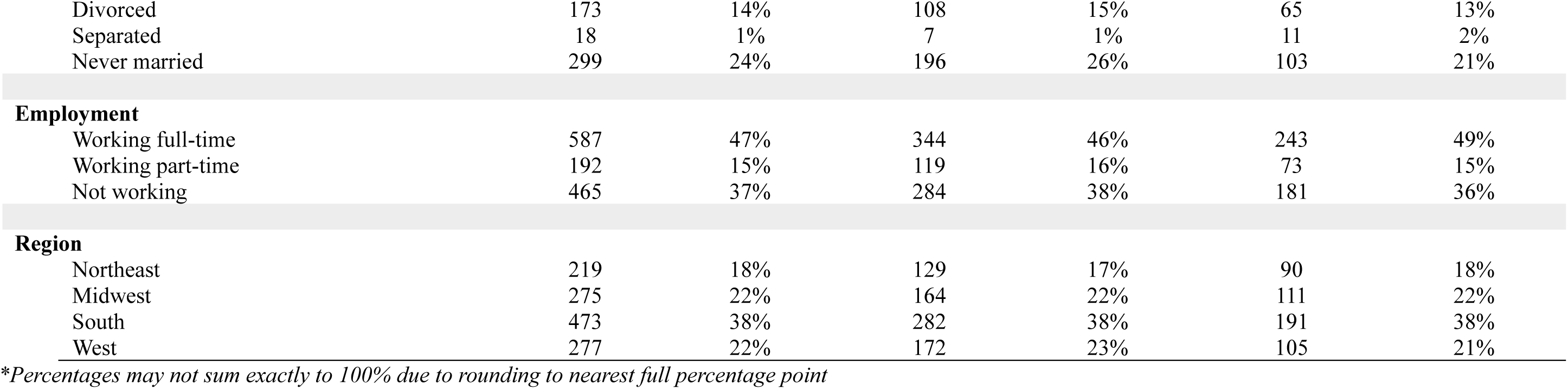
Respondent Demographics.

|  |  | Total<br>(*n*=1,244) |  | Treating with a Product<br>(*n*=747) |  | Not Treating with a Product<br>(*n*=497) |  |
| --- | --- | --- | --- | --- | --- | --- | --- |
|  |  | Mean | Std. Dev. | Mean | Std. Dev. | Mean | Std. Dev. |
| **Age** |  |  |  |  |  |  |  |
|  | Age | 52.5 | 17.0 | 52.1 | 17.1 | 53.2 | 16.8 |
|  |  | N | %* | N | % | N | % |
| **Gender** |  |  |  |  |  |  |  |
|  | Male | 587 | 47% | 341 | 46% | 246 | 50% |
|  | Female | 657 | 53% | 406 | 54% | 251 | 51% |
| **Race/Ethnicity Census Categories** |  |  |  |  |  |  |  |
|  | White, non-Hispanic | 884 | 71% | 544 | 73% | 340 | 68% |
|  | Black, non-Hispanic | 117 | 9% | 66 | 9% | 51 | 10% |
|  | Other, non-Hispanic | 42 | 3% | 22 | 3% | 20 | 4% |
|  | Hispanic | 159 | 13% | 88 | 12% | 71 | 14% |
|  | 2+ Races, Non-Hispanic | 42 | 3% | 27 | 4% | 15 | 3% |
| **Education** |  |  |  |  |  |  |  |
|  | No high school diploma or GED | 64 | 5% | 37 | 5% | 27 | 5% |
|  | High school graduate (high school diploma or the equivalent GED) | 299 | 24% | 176 | 24% | 123 | 25% |
|  | Some college or Associate degree | 353 | 28% | 214 | 29% | 139 | 28% |
|  | Bachelor's degree | 304 | 24% | 191 | 26% | 113 | 23% |
|  | Master's degree or above | 224 | 18% | 129 | 17% | 95 | 19% |
| **Household Income** |  |  |  |  |  |  |  |
| | Under \$10,000 (10) | 35 | 3% | 22 | 3% | 13 | 3% |
| | \$10,000 to \$24,999 (17.5) | 79 | 6% | 42 | 6% | 37 | 7% |
| | \$25,000 to \$49,999 (37.5) | 173 | 14% | 105 | 14% | 68 | 14% |
| | \$50,000 to \$74,999 (62.5) | 183 | 15% | 106 | 14% | 77 | 16% |
| | \$75,000 to \$99,999 (87.5) | 165 | 13% | 105 | 14% | 60 | 12% |
| | \$100,000 to \$149,999 (125) | 245 | 20% | 141 | 19% | 104 | 21% |
| | \$150,000 or more (150) | 364 | 29% | 226 | 30% | 138 | 28% |
| **Marital Status** |  |  |  |  |  |  |  |
|  | Now married | 687 | 55% | 394 | 53% | 293 | 59% |
|  | Widowed | 67 | 5% | 42 | 6% | 25 | 5% |

|  |  |  |  |  |  |  |
| --- | --- | --- | --- | --- | --- | --- |
| Divorced | 173 | 14% | 108 | 15% | 65 | 13% |
| Separated | 18 | 1% | 7 | 1% | 11 | 2% |
| Never married | 299 | 24% | 196 | 26% | 103 | 21% |

Employment
|  |  |  |  |  |  |  |
| --- | --- | --- | --- | --- | --- | --- |
| Working full-time | 587 | 47% | 344 | 46% | 243 | 49% |
| Working part-time | 192 | 15% | 119 | 16% | 73 | 15% |
| Not working | 465 | 37% | 284 | 38% | 181 | 36% |

Region
|  |  |  |  |  |  |  |
| --- | --- | --- | --- | --- | --- | --- |
| Northeast | 219 | 18% | 129 | 17% | 90 | 18% |
| Midwest | 275 | 22% | 164 | 22% | 111 | 22% |
| South | 473 | 38% | 282 | 38% | 191 | 38% |
| West | 277 | 22% | 172 | 23% | 105 | 21% |

Of the total sample, 51% of respondents experienced at least 6 nights of sleep difficulty over the past month, including 33% experiencing insomnia symptoms 11+ nights per month, and 22% experiencing insomnia symptoms 16+ nights per month, as shown in Table 2. Difficulty staying asleep (92%) was most common, followed by difficulty falling asleep (83%) and difficulty waking up too early (83%), as shown in Figure 1. Among those who suffered from more than one sleep issue, difficulty staying asleep was deemed most troublesome by the majority of respondents (53%), as shown in Table 2. 28% of respondents who completed the ISI questionnaire were categorized as suffering from moderate to severe clinical insomnia based on standard index scoring (Table 3).

**Table 2.** Insomnia Symptom Frequency, Evolution, and Most Bothersome.

|  | Total<br>(n=1,244) |  | Treating with a Product<br>(n=747) |  | Not Treating with a Product<br>(n=497) |  |
| --- | --- | --- | --- | --- | --- | --- |
|  | N | % | N | % | N | % |
| **Insomnia Symptom Frequency** |  |  |  |  |  |  |
| None – Because I am successfully treating my difficulty sleeping (taking a medication, supplement, or other substance to aid in sleep) | 75 | 6% | 54 | 7% | 21 | 4% |
| Only one – but I am treating my difficulty sleeping (taking a medication, supplement, or other substance to aid in sleep) | 40 | 3% | 32 | 4% | 8 | 2% |
| 2-5 times (regardless of treatment status) | 511 | 41% | 272 | 36% | 239 | 48% |
| 6-10 times (regardless of treatment status) | 207 | 17% | 118 | 16% | 89 | 18% |
| 11-15 times (regardless of treatment status) | 146 | 12% | 91 | 12% | 55 | 11% |
| 16-20 times (regardless of treatment status) | 94 | 8% | 70 | 9% | 24 | 5% |
| 20+ times (regardless of treatment status) | 171 | 14% | 110 | 15% | 61 | 12% |
| **Evolution of Insomnia Symptoms** |  |  |  |  |  |  |
| This is the first time I have encountered sleep troubles | 63 | 5% | 25 | 3% | 38 | 8% |
| I have sleep troubles occasionally, but sleep troubles are not a common occurrence for me | 298 | 24% | 137 | 18% | 161 | 32% |
| I have recurrent sleep troubles that sometimes appear to go away, only to return | 416 | 33% | 282 | 38% | 134 | 27% |
| My sleep troubles have not gone away since I first started having trouble sleeping | 457 | 37% | 298 | 40% | 159 | 32% |
| Refused | 10 | 1% | 5 | 1% | 5 | 1% |
| **Most Bothersome Insomnia Symptom** |  |  |  |  |  |  |
| Difficulty falling asleep | 212 | 32% | 149 | 35% | 63 | 27% |
| Difficulty staying asleep | 353 | 53% | 221 | 51% | 132 | 57% |
| Problem waking up too early | 98 | 15% | 61 | 14% | 37 | 16% |

\*Percentages may not sum exactly to 100% due to rounding to nearest full percentage point

**Table 3.** ISI Scoring.

|  | Total<br>(n=1,236) |  | Currently Treating<br>(n=742) |  | Not Currently Treating<br>(n=494) |  |
| --- | --- | --- | --- | --- | --- | --- |
|  | N | % | N | % | N | % |
| No Clinically Significant Insomnia | 310 | 25% | 152 | 21% | 158 | 32% |
| Subthreshold Insomnia | 582 | 47% | 349 | 47% | 233 | 47% |
| Clinical Insomnia (Moderate Severity) | 293 | 24% | 208 | 28% | 85 | 17% |
| Clinical Insomnia (Severe) | 51 | 4% | 33 | 4% | 18 | 4% |

*\*Percentages may not sum exactly to 100% due to rounding to nearest full percentage point*

Common impacts of insomnia symptoms include exercising less (51%), eating more (32%), and smoking more (22% of current smokers) as shown in Table 4. Next-day impacts from a poor night of sleep are also notable, including 60% consuming some form of caffeine more often or in a greater amount (compared to typical consumption) due to impacts of poor sleep (Table 4).

**Table 4.** Impacts from Insomnia Symptoms.

|  | Total<br>(*n=Varies*) |  |
| --- | --- | --- |
|  | N | % |
| **Overall Impacts** |  |  |
| I feel my sleep troubles have resulted in me smoking more | 108 | 22% |
| I feel my sleep troubles have resulted in me eating more | 392 | 32% |
| I feel my sleep troubles have resulted in me exercising less | 593 | 51% |
| **What is Done Next Day After Poor Night Sleep** |  |  |
| NET Selected Any Caffeinated Option | 744 | 60% |
| Caffeinated Beverages (i.e., coffee or caffeinated tea, caffeinated soft drinks) | 695 | 56% |
| Caffeine pills (e.g., Vivarin, No-Doze) | 32 | 3% |
| 5-hour energy shots | 27 | 2% |
| Energy drinks (i.e., Red bull, Celsius, Prime, Monster, etc.) | 152 | 12% |
| Prescription medications for alertness (Adderal, Ritalin and Provigil etc.) | 20 | 2% |
| Over-the-counter (OTC) medications for alertness | 15 | 1% |
| Supplements and medicinal herbs like vitamins and Ginseng | 85 | 7% |
| None of the above | 467 | 38% |
| Refused | 3 | 0.2% |

*\*Percentages may not sum exactly to 100% due to rounding to nearest full percentage point*

The majority (60%) of respondents were attempting to treat their insomnia symptoms with some kind of substance. Among those treating within the past three months, the most common were sleep supplements and vitamins (47%), OTC medications for pain and sleep (e.g., Tylenol PM®, Advil PM®, etc.) (25%), and OTC medications for allergies and colds that may result in drowsiness (e.g., Benadryl®, Nyquil^TM^, etc.) (25%). Use of products specifically indicated to address insomnia symptoms was relatively less common, with just 17% of active treaters currently using OTC medications labeled for sleep and 13% utilizing prescription medications prescribed for insomnia symptoms or something similar. Recreational substances were also commonly used, including 21% of treaters utilizing marijuana/cannabis/THC and 14% utilizing alcohol in an attempt to address their insomnia symptoms in the past 3 months. 18% of respondents also reported using prescription medications that were prescribed for another condition as off-label sleep aids. A full breakdown of reported current use can be found in Table 5.

**Table 5.** Treatments Used in Past 3 Months for Insomnia Symptoms.

|  | Total<br>(*n*=1,244) |  |  | Treating with a Product<br>(*n*=747) |  | Not Treating with a Product<br>(*n*=497) |  |
| --- | --- | --- | --- | --- | --- | --- | --- |
|  | N | % | Projectability<br>to US Adult<br>Population | N | % | N | % |
| Regulation of my environmental conditions/surroundings (e.g., bedding, noise machines, room darkening curtains, fans, etc.) | 490 | 39% | 16.5M | 362 | 49% | 128 | 26% |
| Sleep supplements and vitamins (e.g., Melatonin, Magnesium, etc.) | 347 | 28% | 11.8M | 347 | 47% | 0 | 0% |
| Utilization of a bedtime “wind-down” routine | 334 | 27% | 11.4M | 260 | 35% | 74 | 15% |
| Upgrading mattress, linens, or other items for bedroom | 221 | 18% | 7.6M | 168 | 23% | 53 | 11% |
| Over the counter medications for allergy and colds that may result in drowsiness (e.g., Benadryl, Nyquil, store brand allergy and cold relief, etc.) | 188 | 15% | 6.3M | 188 | 25% | 0 | 0% |
| Over the counter medications for pain and sleep (e.g., Tylenol PM, Advil PM, store brand PM pain relief, etc.) | 186 | 15% | 6.3M | 186 | 25% | 0 | 0% |
| Marijuana/cannabis/THC (smoking, vaping, gummies, oils, pills/capsules, and beverages) | 159 | 13% | 5.5M | 159 | 21% | 0 | 0% |
| Prescription medication prescribed for another condition but used to aid in sleep | 132 | 11% | 4.7M | 132 | 18% | 0 | 0% |
| Over the counter medications for sleep (e.g., Unisom, Zzzquil, store brand sleep aid, etc.) | 128 | 10% | 4.2M | 128 | 17% | 0 | 0% |
| Apps/technology designed to help you sleep (e.g., sleep tracker, smart rings, meditation app, etc.) | 126 | 10% | 4.2M | 98 | 13% | 28 | 6% |
| Alcohol | 106 | 9% | 3.8M | 106 | 14% | 0 | 0% |
| Prescription medications for sleep (e.g., Ambien, Lunesta, etc.) | 98 | 8% | 3.7M | 98 | 13% | 0 | 0% |
| CBD/ Cannabidiol (gummies, chews, oils, creams, pills/capsules, and beverages) | 83 | 7% | 3.1M | 83 | 11% | 0 | 0% |
| Cognitive Behavioral Therapy (via technology or in-person with a therapist) | 19 | 2% | 0.8M | 17 | 2% | 2 | 0% |

When sleep sufferers do attempt to treat their insomnia symptoms, they tend to treat frequently. Among those utilizing prescription medications for sleep, marijuana/cannabis/THC, prescription medications for another condition but used to aid in sleep, or sleep supplements and vitamins, median use of these products is daily. Users of alcohol reported treating a median of 5 days per week, and users of CBD/cannabidiol treating a median of 4 days per week (Table 6). Despite the variety and frequency of treatment, perceived benefit was limited or nonexistent. Current treaters reported meeting their personal sleep goal only half of the week. Non-treaters reported a similar median rate of meeting their sleep goals (Figure 2).

**Table 6.** Median Frequency of Use.

| Days per week taking treatment<br>(Median) | Total<br>(*n=varies by option*) |  |
| --- | --- | --- |
|  | N | Median |
| Prescription medications for sleep (e.g., Ambien, Lunesta, etc.) | 75 | 7 |
| Marijuana/cannabis/THC (smoking, vaping, gummies, oils, pills/capsules, and beverages) | 113 | 7 |
| Prescription medication prescribed for another condition but used to aid in sleep | 105 | 7 |
| CBD/ Cannabidiol (gummies, chews, oils, creams, pills/capsules, and beverages) | 42 | 4 |
| OTC meds for allergy and colds that may results in drowsiness (e.g., Benadryl, Nyquil, store brand allergy and cold relief, etc.) | 81 | 2 |
| Sleep supplements and vitamins (e.g., melatonin, magnesium, etc.) | 257 | 7 |
| OTC meds for pain and sleep (e.g., Unisom, Zzzquil, store brand sleep aid, etc.) | 53 | 3 |
| Alcohol | 64 | 5 |

\*Percentages may not sum exactly to 100% due to rounding to nearest full percentage point

**Figure 2.**
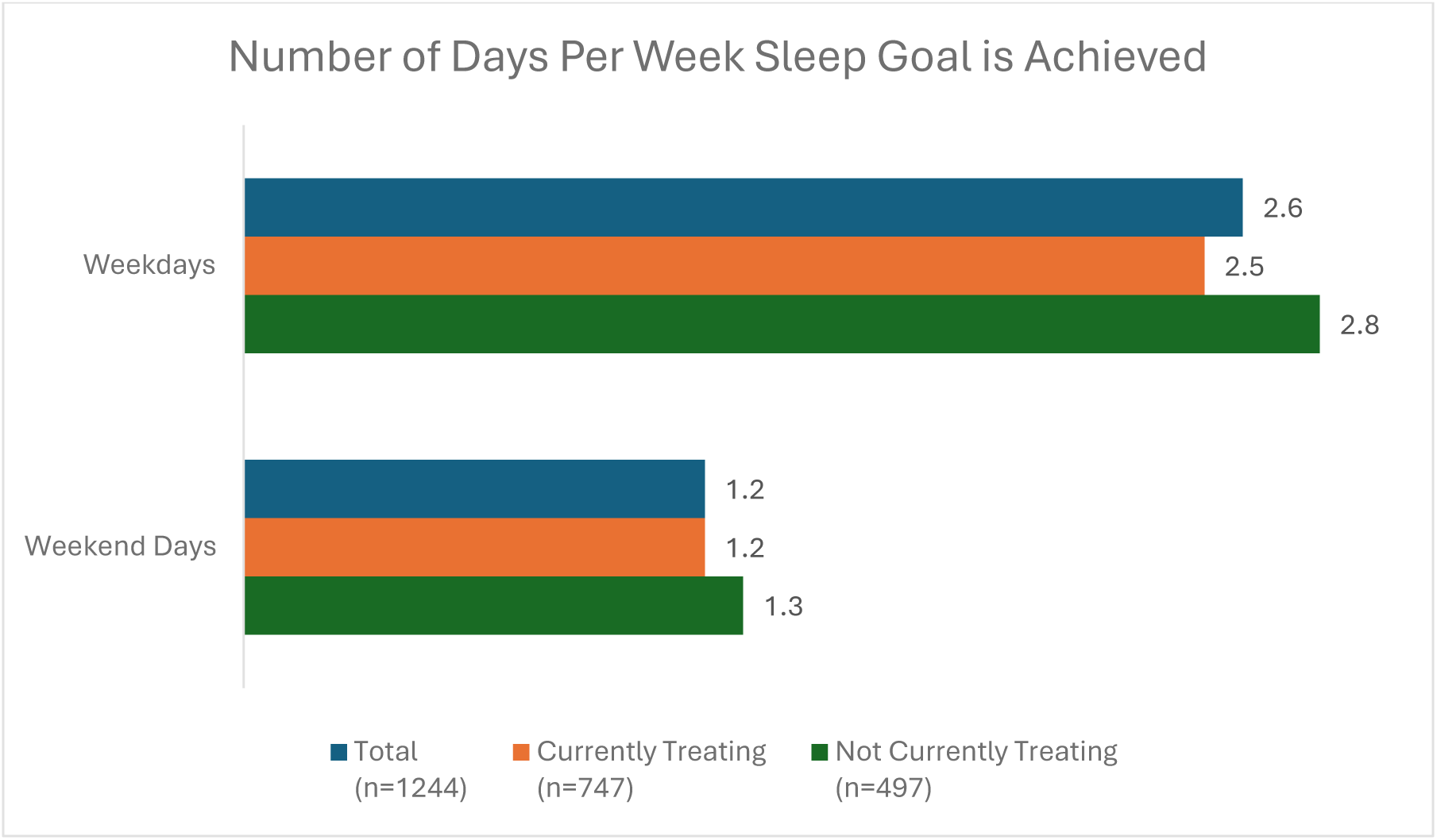
Frequency of Meeting Sleep Goals

Sleep sufferers were open to trying a wide variety of potential sleep treatments. Among those not currently using these treatments, respondents were most willing to try sleep supplements and vitamins (70%), followed by OTC medications for pain and sleep (e.g., Tylenol PM^®^, Advil PM ^®^, etc.) (52%), OTC medications for sleep (52%), OTC medications for allergy and colds that may result in drowsiness (e.g., Benadryl^®^, Nyquil ^TM^, etc.) (45%), prescription medications for sleep (40%), prescription medications prescribed for another condition but used to aid in sleep (38%), CBD/Cannabidiol (35%), marijuana/cannabis/THC (26%), and alcohol (19%). Full distribution can be found in Table 7.

**Table 7.** Openness to Utilizing Other Sleep Treatments.

|  | Total<br>(*n=Varies*) |  | Treating with a Product<br>(*n=Varies*) |  | Not Treating with a Product<br>(*n=Varies*) |  |
| --- | --- | --- | --- | --- | --- | --- |
|  | N | % | N | % | N | % |
| **Sleep supplements and vitamins (e.g., Melatonin, Magnesium, etc.)** |  |  |  |  |  |  |
| NET – At least somewhat open | 681 | 69% | 386 | 79% | 295 | 59% |
| Extremely Open | 170 | 17% | 117 | 24% | 53 | 11% |
| Somewhat open | 511 | 52% | 269 | 55% | 242 | 49% |
| Not open | 295 | 30% | 98 | 20% | 197 | 40% |
| **Over the counter medications for sleep (e.g., Unisom, Zzzquil, store brand sleep aid, etc.)** |  |  |  |  |  |  |
| NET – At least somewhat open | 609 | 51% | 420 | 61% | 189 | 38% |
| Extremely Open | 108 | 9% | 89 | 13% | 19 | 4% |
| Somewhat open | 501 | 42% | 331 | 48% | 170 | 34% |
| Not open | 570 | 48% | 267 | 39% | 303 | 61% |
| **Over the counter medications for pain and sleep (e.g., Tylenol PM, Advil PM, store brand PM pain relief, etc.)** |  |  |  |  |  |  |
| NET – At least somewhat open | 583 | 50% | 366 | 56% | 217 | 44% |
| Extremely Open | 104 | 9% | 74 | 11% | 30 | 6% |
| Somewhat open | 479 | 41% | 292 | 44% | 187 | 38% |
| Not open | 560 | 48% | 285 | 43% | 275 | 55% |
| **Over the counter medications for allergy and colds that may result in drowsiness (e.g., Benadryl, Nyquil, store brand allergy and cold relief, etc.)** |  |  |  |  |  |  |
| NET – At least somewhat open | 516 | 44% | 340 | 51% | 176 | 35% |
| Extremely Open | 94 | 8% | 77 | 12% | 17 | 3% |
| Somewhat open | 422 | 36% | 263 | 40% | 159 | 32% |
| Not open | 636 | 55% | 318 | 48% | 318 | 64% |
| **Cognitive Behavioral Therapy (via technology or in-person with a therapist)** |  |  |  |  |  |  |
| NET – At least somewhat open | 524 | 43% | 338 | 46% | 186 | 38% |
| Extremely Open | 97 | 8% | 60 | 8% | 37 | 8% |
| Somewhat open | 427 | 35% | 278 | 38% | 149 | 30% |
| Not open | 693 | 56% | 387 | 53% | 306 | 62% |
| **Prescription medications for sleep (e.g., Ambien, Lunesta, etc.)** |  |  |  |  |  |  |
| NET – At least somewhat open | 457 | 39% | 315 | 47% | 142 | 29% |
| Extremely Open | 98 | 8% | 81 | 12% | 17 | 3% |
| Somewhat open | 359 | 31% | 234 | 35% | 125 | 25% |
| Not open | 702 | 60% | 352 | 52% | 350 | 70% |
| **Prescription medication prescribed for another condition but used to aid in sleep** |  |  |  |  |  |  |
| NET – At least somewhat open | 426 | 37% | 291 | 45% | 135 | 27% |
| Extremely Open | 58 | 5% | 49 | 8% | 9 | 2% |
| Somewhat open | 368 | 32% | 242 | 38% | 126 | 25% |
| Not open | 701 | 62% | 345 | 54% | 356 | 72% |
| **CBD/ Cannabidiol (gummies, chews, oils, creams, pills/capsules, and beverages)** |  |  |  |  |  |  |
| NET – At least somewhat open | 416 | 35% | 301 | 43% | 115 | 23% |
| Extremely Open | 111 | 9% | 94 | 13% | 17 | 3% |
| Somewhat open | 305 | 25% | 207 | 29% | 98 | 20% |
| Not open | 775 | 65% | 397 | 56% | 378 | 76% |
| **Marijuana/cannabis/THC (smoking, vaping, gummies, oils, pills/capsules, and beverages)** |  |  |  |  |  |  |
| NET – At least somewhat open | 286 | 25% | 205 | 32% | 81 | 16% |
| Extremely Open | 69 | 6% | 55 | 9% | 14 | 3% |
| Somewhat open | 217 | 19% | 150 | 24% | 67 | 14% |
| Not open | 835 | 74% | 423 | 67% | 412 | 83% |
| **Alcohol** |  |  |  |  |  |  |
| NET – At least somewhat open | 209 | 18% | 136 | 20% | 73 | 15% |
| Extremely Open | 19 | 2% | 15 | 2% | 4 | 1% |
| Somewhat open | 190 | 16% | 121 | 18% | 69 | 14% |
| Not open | 956 | 81% | 538 | 79% | 418 | 84% |

*\*Percentages may not sum exactly to 100% due to rounding to nearest full percentage point*

Finally, 74% of respondents indicated that they are not currently seeing an HCP to address their insomnia symptoms, including 48% who had never spoken to an HCP about their sleep (Table 8).

**Table 8.** Healthcare Provider Interactions.

|  | Total<br>(*n*=1,240) |  | No clinically<br>significant insomnia<br>(*n*=310) |  | Subthreshold<br>insomnia (*n*=582) |  | Moderate clinical<br>insomnia<br>(*n*=293) |  | Severe clinical<br>insomnia<br>(*n*=51) |  |
| --- | --- | --- | --- | --- | --- | --- | --- | --- | --- | --- |
|  | N | % | N | % | N | % | N | % | N | % |
| **History of Seeing HCP to Discuss**<br>**Insomnia Symptoms** |  |  |  |  |  |  |  |  |  |  |
| Yes, I currently see a<br>healthcare provider to discuss<br>my trouble sleeping | 328 | 26% | 49 | 16% | 132 | 23% | 118 | 40% | 28 | 55% |
| I do not currently see a<br>healthcare provider to discuss<br>my trouble sleeping but I have<br>in the past | 313 | 25% | 58 | 19% | 168 | 29% | 73 | 25% | 12 | 24% |
| No, I have seen a healthcare<br>provider to discuss my trouble<br>sleeping | 599 | 48% | 202 | 65% | 280 | 48% | 101 | 35% | 11 | 22% |
| **Type of HCP Seen** |  |  |  |  |  |  |  |  |  |  |
| Primary care clinician (e.g.,<br>family doctor, internal<br>medicine, nurse practitioner,<br>physician assistant) | 480 | 75% | 81 | 76% | 224 | 75% | 148 | 78% | 25 | 63% |
| Neurologist | 53 | 8% | 7 | 7% | 23 | 8% | 17 | 9% | 6 | 15% |
| Sleep specialist | 155 | 24% | 22 | 21% | 77 | 26% | 45 | 24% | 10 | 25% |
| Pharmacist | 19 | 3% | 3 | 3% | 9 | 3% | 6 | 3% | 1 | 3% |
| Psychiatrist | 65 | 10% | 8 | 8% | 16 | 5% | 27 | 14% | 14 | 35% |
| Therapist / Psychotherapist | 80 | 13% | 8 | 8% | 29 | 10% | 30 | 16% | 13 | 33% |
| Telehealth provider (i.e., over a<br>phone, video call, or app) | 31 | 5% | 6 | 6% | 9 | 3% | 13 | 7% | 3 | 8% |
| Gynecologist | 21 | 3% | 3 | 3% | 10 | 3% | 6 | 3% | 2 | 5% |
| Other | 31 | 5% | 7 | 7% | 19 | 6% | 4 | 2% | 1 | 3% |
| Refused | 8 | 1% | 1 | 1% | 3 | 1% | 3 | 2% | 0 | 0% |

\*Percentages may not sum exactly to 100% due to rounding to nearest full percentage point

Those with severe clinical insomnia (46%) and moderate clinical insomnia (60%), are also commonly not currently seeing an HCP to address their symptoms. Among those who have seen an HCP, primary care physicians (e.g., family doctors, nurse practitioners, physician assistants, etc.) were most commonly seen (75%). 24% had seen a sleep specialist at some point, 13% had seen a therapist/psychotherapist, and 10% had seen a psychiatrist (Table 8).

## Discussion

The current study provides insight into the prevalence of insomnia symptoms in the US population and the various methods employed by sufferers to address symptoms. Further, this study also provides novel insight into the doctor-patient relationship around sleep. Overall, these results can help guide both future research, health policy and clinical decision making in a coordinated effort to improve the diagnosis and treatment of sleep disturbances.

The study suggests that 58% of American adults suffer two or more nights per month with insomnia symptoms or are actively treating these symptoms (regardless of its frequency or chronicity). This suggests that, at any given time point, approximately 154 million Americans are experiencing insomnia symptoms at least twice per month or are actively managing their ongoing sleep difficulties with medications or other attempted forms of treatment. Difficulty staying asleep was most common among respondents, with most also experiencing comorbid difficulties falling asleep and/or waking up too early. Responses to the validated ISI instrument indicate that approximately 16% of the US adult population, or approximately 42 million Americans, suffer from moderate to severe clinical insomnia. These figures align with broader studies that suggest that approximately 10% of the US population experience symptoms consistent with chronic insomnia, with up to a third reporting some degree of sleep impairment or insomnia [22], further reinforcing the significant and widespread nature of this issue.

With regard to self-care practices, a substantive proportion of people with insomnia symptoms utilized OTC medications for allergies/colds, OTC medications for pain and sleep, marijuana/THC and alcohol. This equates to millions of Americans turning to substances not indicated or labeled for the treatment of insomnia symptoms in an attempt to find something that can potentially offer relief, including approximately 5.5M utilizing cannabis, 4.6M using prescription medication prescribed for something other than sleep, and 3.8M utilizing alcohol. Further, sleep hygiene strategies are among the most commonly turned to methods to address insomnia symptoms, suggesting that this may be a commonly recommended strategy by HCPs despite limited evidence to support their efficacy [23].

These high rates of off-label medication and recreational substance use to aid sleeping warrants concern and further study. This is especially true given the fact that many off-label medications used for sedative purposes (including opioids, anxiolytics, antidepressants, and muscle relaxants) can cause side-effects and may impair functioning, particularly when used at doses and/or for time intervals not intended for the management of insomnia. It should be noted that the reported off-label use of prescription medications in this survey is not expected to account for trazodone but rather includes other classes of prescription medications noted above. Trazodone is likely to be seen as a prescription medication for sleep instead of an off-label prescription through the eyes of a patient.

The vast majority of people with insomnia symptoms were not currently seeing an HCP for their insomnia symptoms, including nearly half who had never seen an HCP for sleep, despite the majority having had insomnia symptoms for a long period of time. Among those who had seen an HCP, most saw a primary care provider. This suggests that many adults that suffer insomnia symptoms are choosing to avoid HCPs. This may be the case for a variety of reasons including poor satisfaction with HCPs, difficulty navigating the health care system due to cost or complexity concerns, or challenges in finding time to seek out an appointment for a condition that could otherwise be dealt with using self-treatment [24–25]. These results suggest that increased availability of evidence-based sleep treatments such as through OTC access could be effective in addressing the needs of adults with insomnia symptoms, especially considering the apparent preference for self-treatment. Increased availability of evidence-based treatments could also prove to be effective in curbing the use of non-evidence-based sleep aids, especially recreational substance use.

### Limitations

The current study possesses many strengths; however, limitations should be noted. This study relied upon self-report surveys. Whereas most cross-sectional surveys face concerns of generalizability due to sample source and composition, recruitment through the rigorously maintained KnowledgePanel^®^ limits generalizability concerns due to the panel’s ability to provide a statistically valid representation of the US population. While standard quantitative survey tactics such as the inclusion of Likert and gradation scales are widely used, there is potential for differing interpretations of scale point meanings from respondent to respondent and thus present a limitation in the interpretation of findings. There is also the potential for under-reporting of off-label medication or recreational substance use due to social stigma and/or legal and confidentiality concerns.

## Conclusions

More than half of American adults reported suffering two or more nights per month of impaired sleep or are actively treating insomnia symptoms. A substantial number were repurposing medications or turning to marijuana/THC and alcohol in an attempt to treat these symptoms. Despite the prevalence of insomnia symptoms, few reported seeking treatments from an HCP.

These results emphasize the widespread nature of sleep difficulties and potential role that a safe and effective OTC sleep aid can play in addressing this public health issue. The current study also highlights that a gap in self-treatment exists for the many millions of adults seeking to treat their symptoms on their own. Critically, improved sleep quality may possess an important public health benefit in the setting of more easily accessible and proven treatments for sleep.

## Data Availability

All data produced in the present study are available upon reasonable request to the authors.

## Writing and Editing Support

Medical writing support was provided by Errol J. Philip, MD, PhD. Molly Atwood, Gregory Smith, Kevin Homler, and Cynthia Herdman provided editorial comments.

## Ethics / Ethics Approval

This study was conducted in full conformance with the Guidelines for GPP published by the International Society of Pharmacoepidemiology (ISPE), the Helsinki Declaration of 1964, and the laws and regulations of the countries in which the research was conducted. Informed consent was collected from all participants prior to participating in the study. No identifiable data was collected during the course of this research or is included in this manuscript. The study (Protocol #2025-0035) was approved by Pearl Institutional Review Board (IRB).

## Author Contributions

Matthew J. Fisher: Conceptualization, Methodology, Writing – Original Draft, Writing – Review & Editing, Supervision

Michael L. Perlis: Conceptualization, Methodology, Writing – Original Draft, Writing – Review & Editing, Supervision

Andrew D. Krystal: Conceptualization, Methodology, Writing – Original Draft, Writing – Review & Editing, Supervision

Matthew J. Heffler: Methodology, Formal analysis, Investigation, Resources, Writing – Original Draft, Writing – Review & Editing, Project administration

## Individual Author Disclosures

Matthew Fisher is an employee and shareholder of Haleon. Haleon manufactures and distributes consumer healthcare products.

Andrew D. Krystal has received research grants from Janssen Pharmaceuticals, Axsome Therapeutics, Attune, Eisai, Harmony, Neurocrine Biosciences, Reveal Biosensors, The Ray and Dagmar Dolby Family Fund, Weill Institute for Neurosciences, and the National Institutes of Health, and consulted for Axsome Therapeutics, AbbVie, Big Health, Eisai, Evecxia, Harmony Biosciences, Idorsia, Janssen Pharmaceuticals, Jazz Pharmaceuticals, Neurocrine Biosciences, Neumora, Neurawell, Otsuka Pharmaceuticals, Sage, Takeda; Stock Options: Neurawell, Big-Health.

Michael L. Perlis has received research grants from Teva, Cephalon, Sanofi-Aventis, Axsome Therapeutics, consulted for Guidepoint and Gerson-Lehman Group, and has been a clinical advisor for Sanofi-Aventis, Takeda, Sepracor, Actelion, SleepEasily, InsomniSolv, Scientific Software Technologies, Lumosity, Nexalin, Anavex, PlayPower, NeuroUx, Z-Brain, Haleon/GSK.

Matthew J. Heffler is an employee of Ipsos-Insight, LLC, who was contracted to conduct this study on behalf of Haleon.

