## Supplemental Table 1 for "The State of Sleep: Understanding Insomnia Symptoms and Treatments in US Adults"

### Supplementary Table 1. Weighting Benchmark Distributions

| | *Consented respondents* | | *18+ US Population Benchmarks** | *Qualified respondents* | |
|---|---|---|---|---|---|
| | *Un-weighted* | *Weighted* | | *Weighted* | *Un-weighted* |
| | **Percent** | **Percent** | **Percent** | **Percent** | **Percent** |
| **18-29 Male** | 7.51 | 10.02 | **10.02** | 6.75 | 5.14 |
| **18-29 Female** | 7.6 | 9.81 | **9.81** | 8.53 | 6.67 |
| **30-44 Male** | 13.02 | 13.05 | **13.05** | 12.53 | 11.98 |
| **30-44 Female** | 10.97 | 13.01 | **13.01** | 13.62 | 11.33 |
| **45-59 Male** | 11.64 | 11.45 | **11.45** | 11.86 | 11.82 |
| **45-59 Female** | 10.75 | 11.72 | **11.72** | 14.07 | 12.78 |
| **60+ Male** | 18.75 | 14.34 | **14.34** | 14.25 | 18.25 |
| **60+ Female** | 19.77 | 16.6 | **16.6** | 18.37 | 22.03 |
| | **Percent** | **Percent** | **Percent** | **Percent** | **Percent** |
| **White, Non-Hispanic** | 66.55 | 60.76 | **60.76** | 66.1 | 71.06 |
| **Black, Non-Hispanic** | 10.53 | 12.08 | **12.08** | 11.25 | 9.41 |
| **Other, Non-Hispanic** | 4.98 | 7.64 | **7.64** | 4.86 | 3.38 |
| **Hispanic** | 14.39 | 17.88 | **17.88** | 16.13 | 12.78 |
| **2+ Races, Non-Hispanic** | 3.55 | 1.64 | **1.64** | 1.66 | 3.38 |
| | **Percent** | **Percent** | **Percent** | **Percent** | **Percent** |
| **Northeast Metro** | 16.44 | 15.92 | **15.92** | 15.58 | 16.08 |
| **Northeast Non-metro** | 1.51 | 1.28 | **1.28** | 1.42 | 1.53 |
| **Midwest Metro** | 16.57 | 16.3 | **16.3** | 17.11 | 17.12 |
| **Midwest Non-metro** | 4.62 | 4.15 | **4.15** | 4.62 | 4.98 |
| **South Metro** | 32.07 | 33.09 | **33.09** | 32.75 | 31.59 |
| **South Non-metro** | 5.78 | 5.6 | **5.6** | 6.19 | 6.43 |
| **West Metro** | 21.28 | 21.37 | **21.37** | 20.44 | 20.74 |

| | | | | | |
|---|---|---|---|---|---|
| **West Non-metro** | 1.73 | 2.29 | **2.29** | 1.89 | 1.53 |

| | **Percent** | **Percent** | **Percent** | **Percent** | **Percent** |
|---|---|---|---|---|---|
| **Less than HS** | 5.86 | 9.26 | **9.26** | 7.97 | 5.14 |
| **HS** | 25.28 | 28.66 | **28.66** | 27.54 | 24.04 |
| **Some college** | 27.23 | 26.29 | **26.29** | 27.51 | 28.38 |
| **Bachelor or higher** | 41.63 | 35.79 | **35.79** | 36.99 | 42.44 |

| | **Percent** | **Percent** | **Percent** | **Percent** | **Percent** |
|---|---|---|---|---|---|
| **Under $25,000** | 9.55 | 9.8 | **9.8** | 9.02 | 9.16 |
| **$25,000-$49,999** | 13.46 | 14.3 | **14.3** | 14.18 | 13.91 |
| **$50,000-$74,999** | 14.22 | 14.71 | **14.71** | 15.51 | 14.71 |
| **$75,000-$99,999** | 13.06 | 12.52 | **12.52** | 13.1 | 13.26 |
| **$100,000-$149,999** | 19.24 | 19.28 | **19.28** | 20.32 | 19.69 |
| **$150,000 and over** | 30.48 | 29.38 | **29.38** | 27.88 | 29.26 |
